# Walking speed and propulsion asymmetry as distinct diagnostic biomarkers of post-stroke gait impairment: Towards precision rehabilitation after stroke

**DOI:** 10.64898/2026.09.14.26360742

**Authors:** Dheepak Arumukhom Revi, Johanna Spangler, Ariearavanan Chinnappan, Stefano M.M. De. Rossi, Mark G. Bowden, Terry D. Ellis, Louis N. Awad

**Affiliations:** Sargent College of Health and Rehabilitation Sciences, Boston University, Boston, MA; Department of Mechanical Engineering, Boston University, Boston, MA; Division of Clinical Integration and Research, Brooks Rehabilitation, Jacksonville, FL

**Author notes:** **Corresponding Authors:** Dheepak Arumukhom Revi and Louis Awad.

**Keywords:** stroke, propulsion asymmetry, precision rehabilitation, movement phenotypes, diagnostic biomarkers

## Abstract

Stroke-induced hemiparesis often results in slow walking and an asymmetrical gait; however, these impairments vary widely across patients. Indeed, some individuals walk fast despite severe gait asymmetry, whereas others walk slowly despite mild asymmetry. We posit that speed and symmetry reflect distinct dimensions of walking and may serve as complementary biomarkers of stroke recovery. In this cross-sectional study, 57 individuals with chronic hemiparesis were classified as “slow” or “fast” and as having “severe” or “mild” asymmetry, using pre-defined thresholds (i.e., speed = 0.8 m/s; symmetry = 31.5%). Kruskal-Wallis and regression analyses were used to evaluate differences in gait biomechanics (i.e., limb and joint propulsive power and the metabolic cost of walking) and ambulatory function (i.e., speed, distance, and functional balance) associated with these impairment classifications. When examined independently, slower speeds and greater asymmetry were similarly associated with a higher metabolic cost of walking (Δ range: 77% to 83% higher, p < 0.05) and reduced balance and walking function (Δ range: 26% to 82% lower, p < 0.05). However, the two classification approaches revealed different underlying biomechanical mechanisms: whereas slower walking corresponded to reduced paretic propulsive power (Δ: 71% lower, p < 0.05), greater asymmetry indicated a distal-to-proximal shift in power generation (Δ: 200% shift from the ankle to the hip, p < 0.05). Furthermore, multiple regression analyses revealed that each classifier was an independent predictor of metabolic cost (R²=0.70, p < 0.001) and ambulatory function (R² range: 0.34 to 0.55, p < 0.001). These findings demonstrate the complementary diagnostic value of measuring *both* walking speed and propulsion asymmetry, supporting the development of multi-dimensional phenotypes to enable precision neurorehabilitation.

## Introduction

Stroke is a leading cause of long-term disability, affecting nearly 100 million people worldwide [Feigin 2021]. Post-stroke neuromotor impairments are diverse but commonly result in a slow, asymmetric, inefficient, and unstable gait [Reisman 2009, Farris 2015, Combs 2013, Mahon 2015]. Among the key contributors to these impairments—and an established diagnostic biomarker of hemiparetic severity—is the reduced ability of the paretic limb to generate forward propulsion [Bowden 2006, Roelker 2019]. In neurologically intact individuals, symmetrical propulsion from both limbs enables efficient step-to-step acceleration of the body [Kuo 2010]. In contrast, poststroke neuromotor deficits impair the paretic limb’s ability to generate propulsion [Bowden 2006], leading to significant gait propulsion asymmetry and slower walking speeds. Although the functional and metabolic consequences of impaired paretic propulsion have been explored [Bowden 2008, Kuo 2010, Reisman 2009], it is unclear whether propulsion asymmetry and slow walking speed share similar underlying biomechanical mechanisms and contributions to deficits in balance and walking function, or if they reflect distinct mechanisms and contributions. Differentiating how deficits in propulsion asymmetry and walking speed are mechanistically and functionally related is critical for advancing the field of post-stroke gait rehabilitation, and specifically the subareas of precision diagnostics and targeted interventions.

### Biomarkers of functional recovery

While walking speed is a widely recognized measure of walking ability [Fritz 2009, Fulk 2010, Grau-Pellicer 2019, Fulk 2017], propulsion asymmetry has emerged as a critical indicator of walking performance [Mahon 2015, Barroso 2017, Bowden 2006], a predictor of gait recovery [Awad 2016], and a target for therapeutic interventions [Hsiao 2016, Awad 2016, Porciuncula 2021, Swaminathan 2023, Porciuncula 2023, Parikh 2026]. Although propulsion and walking speed are biomechanically coupled [Kuo 2005], diverging patterns of impairment across individuals poststroke suggest that these metrics reflect different dimensions of functional recovery and may provide unique diagnostic information on underlying mechanisms of impairment. Indeed, a notable and seemingly paradoxical clinical observation is that individuals with severe propulsion asymmetry may walk at relatively fast speeds, while others with mild asymmetry walk slowly [Bowden 2006]. Thus, we hypothesize that propulsion asymmetry and walking speed are distinct yet complementary biomarkers of functional recovery and predict that they will independently explain the variation in the recovery of functional balance and walking function observed after stroke.

### Biomarkers of biomechanical mechanisms

During neurotypical walking, positive work is cumulatively generated by the muscles of the trailing limb to produce an anteriorly directed (i.e., forward propulsive) ground reaction force. Inter-limb coordination enables the symmetrical repeating of this process by the other limb during its stance phase, facilitating the metabolically efficient acceleration typically associated with human bipedal walking [Kuo 2005]. In post-stroke hemiparesis, this coordination is disrupted, with the paretic limb generating less positive work while the total work completed by both limbs increases; consequently, post-stroke gait is highly metabolically inefficient [Kuo 2010, Farris 2015, Jonkers 2009]. Furthermore, when deepening the analysis to the joint level, post-stroke hemiparesis is associated with heterogeneous deficits in the intra-limb coordination of power generation [Sawicki 2009, Farris 2015], including a proportionate reduction from each joint to a redistribution across joints. Indeed, individuals poststroke generate, on average, less power from the ankle [Farris 2015, Jonkers 2009], and in some individuals, this is compensated by increased work by the hip muscles [Farris 2015]. This distal-to-proximal (i.e., ankle-to-hip) redistribution of mechanical power is consistent with previous reports of plantarflexor weakness compensated by the hip flexors [Nadeau 1999]. Because different joints have different elastic energy storage and return mechanisms during walking [Sawicki 2009, Alexander1988], compensatory power redistribution from the more efficient ankle muscles to the less efficient hip muscles [Alexander1988] is thought to also contribute to the higher metabolic cost of walking after stroke.

Although slow walking is also associated with a metabolically inefficient gait [Donelan 2001, Ellis 2013], the mechanisms are thought to be different and include suboptimal utilization of tendon elasticity for energy recycling [Ellis 2013]. Indeed, although reduced total power generation is biomechanically expected at slower speeds [Donelan 2001, Donelan 2002], slow walking after stroke is not necessarily associated with the impaired power generation strategies observed in asymmetric individuals—i.e., a redistribution of power generation across limbs or, when examined within limbs, across joints. Thus, we hypothesize that different patterns of power generation impairment underlie the higher metabolic cost of walking observed in persons with propulsion asymmetry and persons with slow walking speed. Consequently, we predict that walking speed and propulsion asymmetry will independently explain the higher metabolic cost of walking observed in people poststroke.

### Advancing precision rehabilitation

The objective of this study is to determine if propulsion asymmetry and walking speed may be complementary biomarkers that explain post-stroke variation in biomechanical mechanisms of power generation, the metabolic cost of walking, and functional recovery—including walking endurance (6-Minute Walk Test), functional balance (Functional Gait Assessment), and functional mobility (Timed-Up and Go). The findings of this study will elucidate the diagnostic value of including both walking speed and propulsion asymmetry as part of multi-dimensional phenotypes with potential to advance precision rehabilitation that delivers “the right intervention, for the right individual, at the right time” [French 2022].

## Methods

This paper compiles data prospectively collected for two distinct studies of biomechanical (study 1) and metabolic (study 2) outcomes, which shared an overlapping subset of post-stroke and healthy control study participants **(Figure 1)**. The objective of study 1 was to determine if classifying individuals with poststroke hemiparesis based on their propulsion asymmetry and walking speed reveals group differences in power generation impairment. Study 2 builds on these findings by evaluating if the differences in power generation impairment observed across these classification groups propagate to differences in the metabolic cost of walking. Ambulatory function outcomes collected during both studies were supplemented with laboratory and publicly available normative values (where available) to provide a unique multimodal dataset suited to the objectives of this paper.

**Figure 1:**
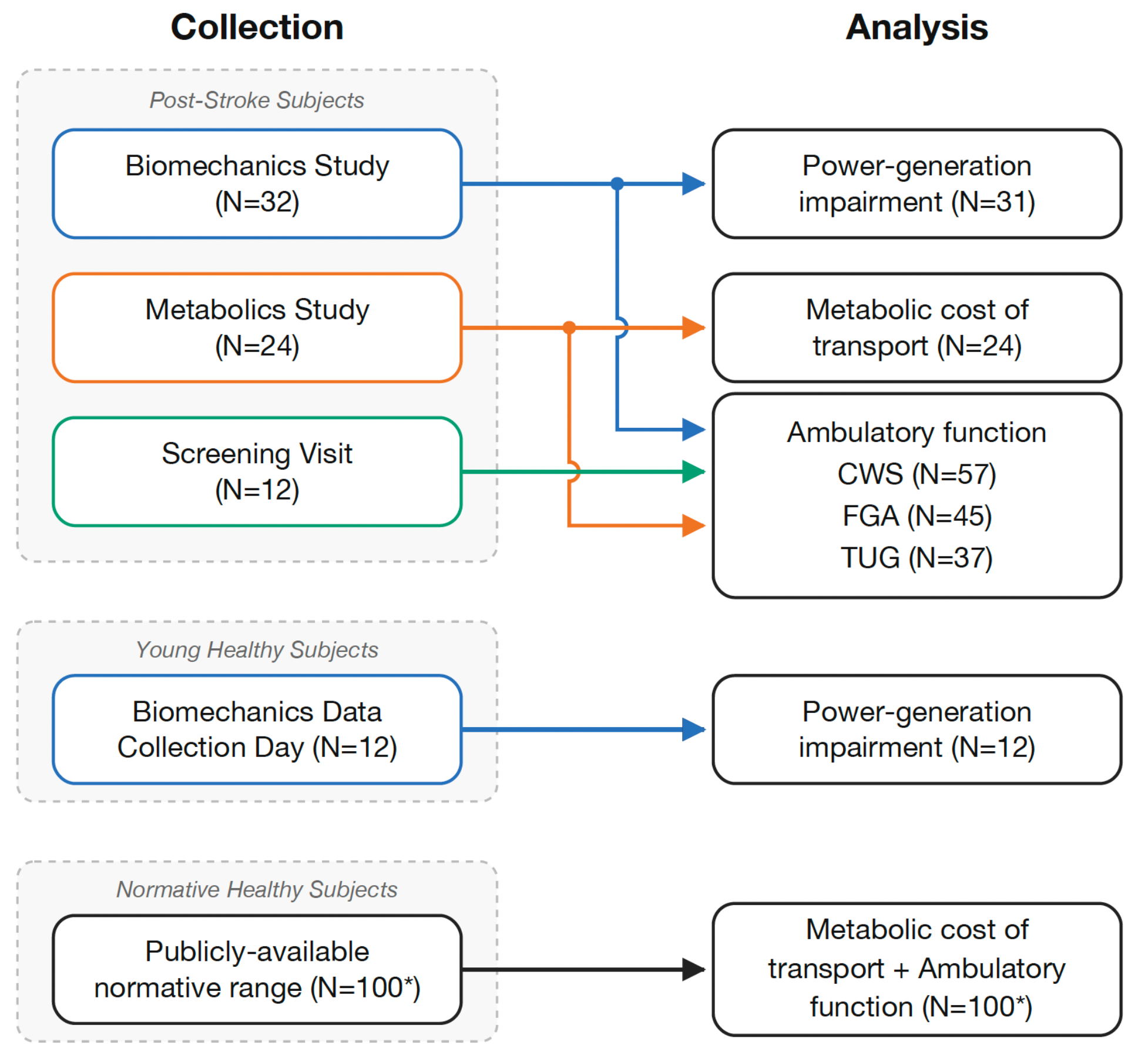
Data collection and analysis flow; Abbreviations: CWS = Comfortable walking speed, FGA = Functional Gait Assessment, TUG = Time Up and Go. Publicly available normative range includes the following studies [Morbach 2025, Bohannon 2011, Walker 2007, Bohannon 2006, Berryman 2012] where averages and standard deviation of the set collected on up to 2762 individuals were used; *100 normative individual data points was estimated (See *Normative healthy control* section)

### Participants

Data from 57 unique individuals in the chronic phase after stroke (sex: 41M/16F, age: 60.0 ± 10.4 years, height: 172 ± 9 cm, weight: 86.7 ± 19.9 kg, paresis: 32L/25R, onset: 6.3 ± 5.6 years) and twelve young healthy control subjects (gender: 6M/6F age: 25.6 ± 3.7 years, height: 172 ± 12 cm, weight: 68.3 ± 16.4 kg) were included in this study. Study size was determined by the number of participants available from the contributing studies. All study procedures were approved by the Institutional Review Board at Boston University. Participants were recruited from the community in the greater Boston area.

The inclusion criteria for study participants who were post-stroke consisted of being greater than six months post-stroke, having the ability to walk without the assistance of another individual. The exclusion criteria included other comorbidities that impaired walking ability, resting heart rate outside the range of 40 to 100 beats per minute, resting blood pressure outside the range of 90/60 to 170/90 mmHg, and the inability to communicate with the investigators.

### Study procedures

#### Biomechanical data collection

A subset of thirty-two individuals with post-stroke hemiparesis (sex: 26M/7F, age: 58.8 ± 10.2 years, height: 173 ± 9 cm, weight: 86.3 ± 18.9 kg, paresis: 16L/16R, stroke onset: 6.4 ± 4.8 years) and all twelve healthy controls (gender: 6M/6F age: 25.6 ± 3.7 years, height: 172 ± 12 cm, weight: 68.3 ± 16.4 kg) participated in biomechanics data collection procedures between September 2019 and June 2022. A fully instrumented 6-minute walk test (6MWT) was performed around a 26.6 m oval indoor track consisting of two 10-meter straightaways separated by 3.3m turns on either end. Instrumentation included: motion capture cameras to track retro-reflective markers placed on the lower limb (200Hz, 18-camera motion-capture, Qualisys, Göteborg, Sweden), wireless internal measurement units placed on the pelvis, shank, and thigh (100Hz, MTw Awinda, Xsens, Enschede, Netherlands), and six forceplates (2000Hz, Bertec, Columbus, OH) embedded in the walkway. Together, these instruments respectively enabled concurrent collection of gait kinematic, inertial, and kinetic signals **(Figure 2)**. More specifically, while inertial data were collected throughout the entire walkway, one of the two 10-meter straightaways included the capture area used by the motion capture and forceplates. All signals were time-synced using a synchronization pulse triggered at the start of data collection and were filtered using a zero-lag 4^th^ order low pass Butterworth filter with a 10 Hz cutoff frequency. A static trial was collected before starting the 6MWT to serve as a baseline reference for the kinematic and inertial data.

**Figure 2:**
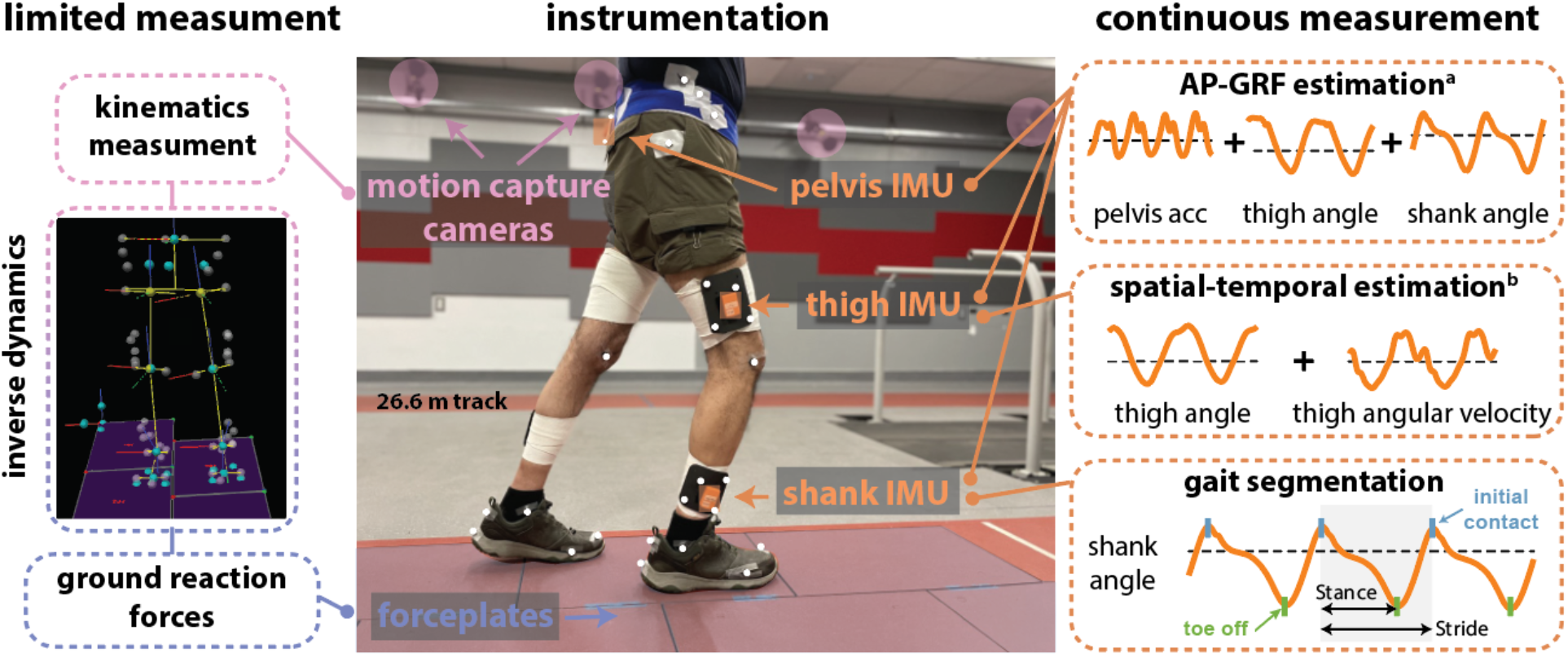
Lab Instrumentation Setup. Motion capture and forceplate data were used to perform inverse dynamics calculations; these data are limited to when forceplate data are available. In addition, we utilized IMU gait estimation algorithms that we previously developed and validated to enable estimation of gait propulsion^a^ [ArumukhomRevi 2020] and spatial-temporal parameters^b^ [ArumukhomRevi 2021] on a stride-by-stride basis throughout the entire 6-minute walk test.

#### Metabolic data collection

A subset of twenty-four individuals (sex: 18M/6F, age: 58.8 ± 10.7 years, height: 173 ± 8 cm, weight: 86.7 ± 18.8 kg, paresis: 14L/10R, onset: 6.6 ± 4.2 years) participated in metabolic data collection procedures between September and December 2023. In the same 26.6m oval indoor track as used for the biomechanics data collection, study participants performed the 6MWT while wearing an indirect calorimetry system (COSMED, K5, Italy). The six embedded forceplates (2000Hz, Bertec, Columbus, OH) were used to collect ground reaction force data during the 6MWT. Prior to the 6MWT, participants completed a five-minute quiet standing period to establish a resting metabolic data. Net metabolic cost of transport was computed by subtracting the average resting metabolic during quiet standing from the gross metabolic average of the last two minutes of walking, then dividing by average walking speed.

#### Ambulatory function data collection

All 57 individuals post-stroke completed a 10-meter walk test at a comfortable walking speed (CWS) and the instrumented 6MWT. A subset of 45 individuals also completed the Functional Gait Assessment (FGA) [Wrisley 2004, Walker 2007], and a subset of 37 individuals completed the Timed Up and Go (TUG) test [Podsiadlo 1991, Bohannon 2006]. All participants were recruited and data collected between September 2019 and June 2024.

### Data Processing and Analysis

#### Inverse dynamics analysis

Kinematic and kinetic biomechanical analyses were conducted using all biomechanical data collected during steps with forceplate data. Ankle, knee, and hip power were computed using the Visual 3D software (C-Motion, Germantown, MD USA) using the default segment geometries and six degrees of freedom between all segments **(Figure 2)**. The average positive power for each joint was calculated using the trapezoid method, where all the positive power during the gait cycle was added and divided by stride time [Farris 2012]. The total average positive power was defined as the combined average positive power of the hip, knee, and ankle [Farris 2012]. Percent positive joint power was defined as positive power from a specific joint (ankle, knee, or hip) divided by the total average positive power. To quantify the distribution of joint power, the ankle-to-hip ratio (A-H ratio) was calculated and defined as total positive ankle power divided by total positive hip power, where:

• A-H ratio = 1 indicates equal contribution from distal (ankle) and proximal (hip) joints

• A-H ratio < 1 indicates more proximal power (i.e., more hip than ankle power), and

• A-H ratio > 1 indicates more distal power (i.e., more ankle than hip power).

In healthy controls, data from the right limb was used by default as substantive symmetry across limbs was assumed [Sadeghi 2000].

#### Inertial data measurement during the 6MWT

Five inertial measurement units (IMUs) were securely attached to marker clusters, with two IMUs located laterally on each shank, two IMUs located laterally on each thigh, and one IMU located posteriorly on the pelvis [ArumukhomRevi 2020]. All IMUs were positioned such that one of the IMU axes always moved along the sagittal plane of motion. The shank IMU was used to define each gait cycle [ArumukhomRevi 2020]. Using the estimation methods described in previous work, anterior-posterior ground reaction forces (AP-GRF) were estimated, from which propulsion metrics were extracted [ArumukhomRevi 2020], and spatial-temporal measures [ArumukhomRevi 2021] were estimated bilaterally for every stride taken during the 6MWT. Data collected during strides with available forceplate strikes were used to calibrate the estimation algorithms **(Figure 2)**.

### Normative healthy control

Publicly available normative range that included participants with similar characteristics as our subjects (e.g., age, height, speed) were obtained from the literature to serve as a healthy control comparator for our functional and metabolic data analyses. Specifically, a normative 6MWT distance of 570 m [451 m – 694 m] was obtained from Morbach et al. [Morbach 2025] based on a sample of 2,762 healthy individuals with an average age of 70 years and an average height of 170 cm. A normative comfortable walking speed value of 1.34 m/s [1.26 m/s – 1.41 m/s] was obtained from Bohannon et al. [Bohannon 2011] based on a sample of 941 male individuals with an age between 60-70 years. A normative FGA score of 27.1 ± 2.3 points was obtained from Walker et al. [Walker 2007] based on a sample of 63 individuals with an age between 60-70 years. A normative TUG time of 8.1 sec [7.1 sec to 9 sec] was obtained from Bohannon et al. [Bohannon 2006] based on a sample of 176 individuals with an age between 60-70 years. A normative net metabolic cost of walking of 0.140 ± 0.029 mL/kg/m was obtained from Berryman et al. [Berryman 2012] based on a sample of 20 individuals with an average age of 69 years, walking overground with an average speed of 0.89 m/s. For all normative healthy studies not collected in our lab, individual subject data were not published as part of the respective studies. Therefore, we created a test dataset of 100 randomly selected datapoints using a bootstrap approach such that the normative mean and standard deviations reported in the respective studies were recreated.

To the best of our knowledge, no age-matched normative values are available for propulsion asymmetry, especially with regards to older adults. While aging does lead to a decline in power generation capacity and can lead to a hip-dominant walking strategy [Boyer 2023], it is not likely that a significant asymmetry would emerge across limbs due to aging alone [Farris 2015, Sadeghi 2000, Cofre 2011], unlike in individuals with post-stroke hemiparesis. Therefore, the propulsion asymmetry values computed from the data collected with our healthy young cohort are used in all healthy comparisons.

### Analysis

#### Speed-based classification

For all study participants, walking speed was calculated either (1) from the distance walked during the six-minute walk test divided by the 360 seconds required to complete the test, or (2) if IMU data were available, as the average speed across strides walked during the test [ArumukhomRevi 2021]. Our prior work demonstrates the equivalency of these approaches [ArumukhomRevi 2021]. To classify individuals based on their walking speed, we used the long-distance walking speed threshold reported by Fulk et al. [Fulk 2017], where individuals walking less than 288m during the 6MWT (i.e., slower than 0.80 m/s) were reported to more likely be limited community ambulators (i.e., walking less than 7500 steps/day) and individuals walking more than 288m during the 6MWT (i.e., faster than 0.80 m/s) were reported to more likely be unlimited community ambulators (i.e., walking greater than 7500 steps/day). For the purposes of our study, individuals with a 6MWT distance less than 288m were classified as “slow”, while individuals with a 6MWT distance greater than 288m were classified as “fast”.

#### Propulsion-based classification

Propulsion asymmetry was defined as the positive paretic propulsive impulse divided by the sum of the positive paretic and non-paretic impulses, such that 50% indicates perfect symmetry [Bowden 2006]. The propulsive impulse generated by each limb was measured directly from forceplates and, when IMU data were available, estimated using a high-accuracy estimation algorithm previously described [ArumukhomRevi 2020], and averaged over the entirety of the 6MWT. To classify individuals based on their propulsion asymmetry, we computed a propulsion asymmetry threshold based on prior work by Bowden et al. [Bowden 2006], where individuals with mild, moderate, and severe hemiparetic severity were reported to have mean propulsion asymmetry values of 49%, 36%, and 16% respectively. To identify a robust propulsion asymmetry threshold, we conducted an ROC analysis using the propulsion and clinical data (Brunnstrom Stages) contained in the dataset published by Bowden et all (see **Appendix A**). We found that a propulsion asymmetry threshold of 31.5% could distinguish individuals with moderate-to-severe hemiparetic severity from individuals with mild hemiparetic severity. Thus, for the purposes of our study, individuals with a median propulsion asymmetry less than 31.5% were classified as having “moderate-severe” hemiparesis, while individuals with a median propulsion asymmetry greater than 31.5% were classified as having “mild” hemiparesis.

### Statistical analysis

Group differences were analyzed using a Kruskal-Wallis test, with alpha set to 0.05. If the group level data were significant, multiple Wilcoxon rank sum tests were performed, with post-hoc correction using the Holm-Sidak method. All data analyses were performed in MATLAB R2021a (MATHWORKS, Natick, MA) with available cases for each outcome. Individuals with greater paretic propulsion than non-paretic propulsion (Pp>52.5%, allowing for a small 2.5% asymmetry) were excluded in our analysis due to the possibility of different neurological control strategies [Bowden 2012]. To assess the independent contributions of the walking speed and propulsion-based classifications to the variance in metabolic and ambulatory function outcomes, we conducted multiple linear regression analyses and compared the standardized beta coefficients for the individual predictors. In addition, the interaction between the classifiers was tested and, if significant, included in the final model.

## Results

### Biomechanical assessment of power generation

Of the 32 individuals poststroke that completed biomechanical assessments, one was excluded from all analyses due to having a higher contribution to propulsion from the non-paretic limb compared to the paretic limb, which indicates a different neuromotor control strategy [Bowden 2006]. Of the remaining 31 individuals, 13 were classified as slow and the remaining 18 were classified as fast based on the speed-based classification cut-off of 0.80 m/s. Based on the propulsion-based classification cut-off of 31.5%, 14 were classified as having moderate-severe propulsion deficits and the remaining 17 were classified as having mild propulsion deficits.

#### Speed-based classifier

As expected, when compared to healthy controls, individuals poststroke in both the fast and slow classification groups had significantly lower total positive power generation by their paretic and non-paretic limbs (p<0.001). While the total positive power generated was significantly higher in the fast versus slow groups (p<0.001), the relative contribution of ankle and hip power to total power (i.e., A-H ratio) was not significantly different for either the paretic or non-paretic limbs (p>0.05) (see **Table 1**, **Figure 3A**).

**Table 1:** Positive power generation across joints based on speed classification.

| | Slow group<br>($<0.80$ m/s) | Fast group<br>($>0.80$ m/s) | Healthy<br>control | K-W<br>p value | Post-hoc<br>sig pair |
| --- | --- | --- | --- | --- | --- |
| N | 13 | 18 | 12 | - | - |
| Pp (%) | 30.84 | 36.08 | 48.81 | $<0.001^*$ | 2-3,1-3 |
| ***Paretic Limb Metrics*** |  |  |  |  |  |
| Total power (J/kg) | 0.26 (0.26) | 0.98 (0.66) | 2.42 (0.68) | $<0.001^*$ | 1-2, 2-3,<br>1-3 |
| Ankle power (%) | 17.4 (21.1) | 21.9 (11.2) | 26.9 (7.1) | 0.16 | - |
| Knee power (%) | 25.9 (7.8) | 19.7 (16.3) | 25.0 (5.5) | 0.37 | - |
| Hip power (%) | 50.4 (22.3) | 54.0 (25.0) | 48.5 (9.7) | 0.40 | - |
| A/H ratio | 0.30 (0.55) | 0.44 (0.42) | 0.56 (0.26) | 0.37 | - |
| ***Non-Paretic Limb Metrics*** |  |  |  |  |  |
| Total power (J/kg) | 0.36 (0.20) | 1.25 (0.86) | 2.42 (0.68) | $<0.001^*$ | 1-2, 2-3,<br>1-3 |
| Ankle power (%) | 28.4 (13.8) | 29.1 (10.2) | 26.9 (7.1) | 0.57 | - |
| Knee power (%) | 15.5 (12.6) | 14.1 (7.2) | 25.1 (5.5) | $0.005^*$ | 2-3, 1-3 |
| Hip power (%) | 47.5 (26.0) | 55.2 (13.2) | 48.5 (9.7) | 0.17 | - |
| A/H ratio | 0.54 (0.53) | 0.59 (0.38) | 0.56 (0.26) | 0.69 | - |

**Figure 3:**
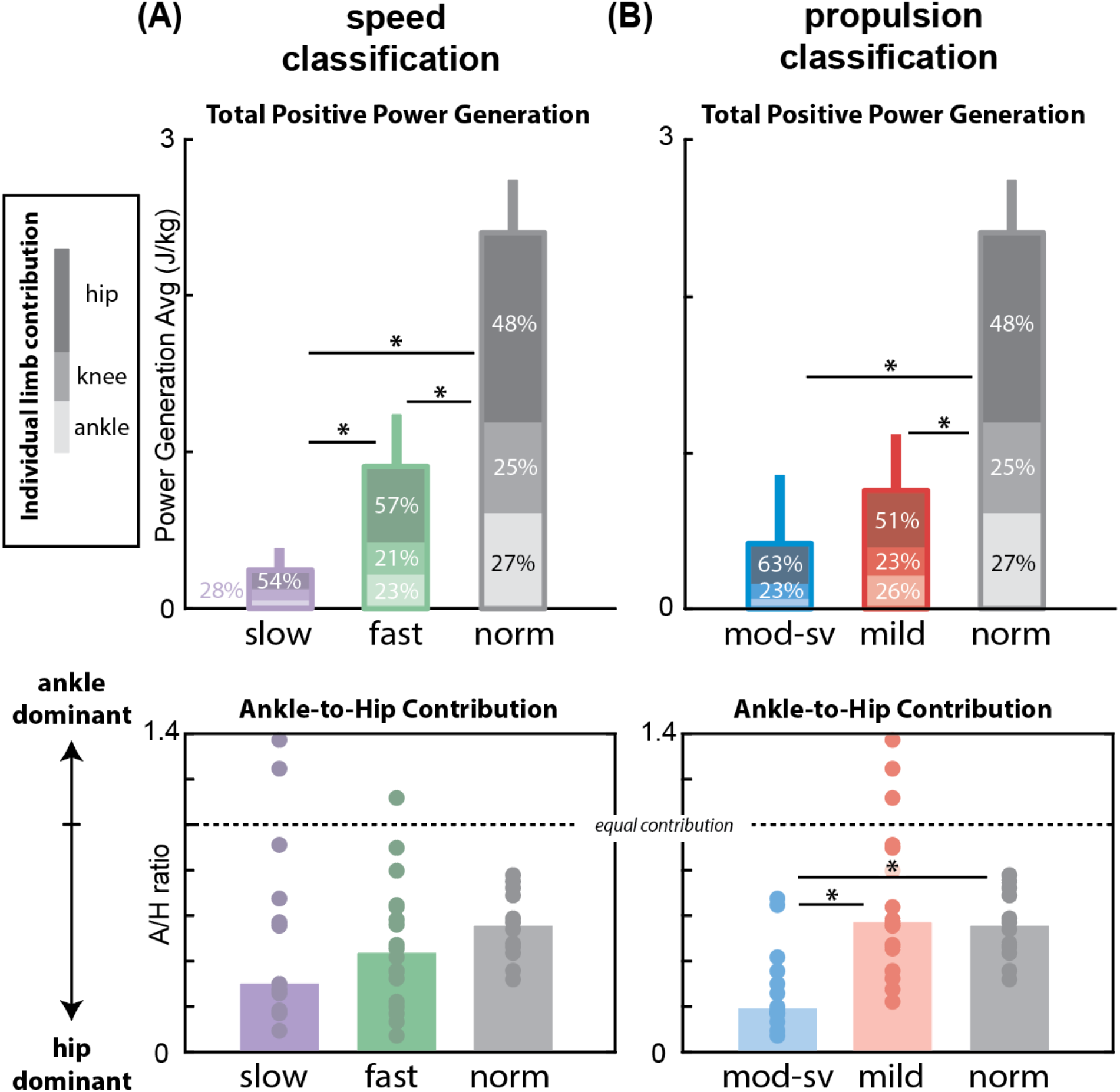
Power distribution across speed and propulsion classification groups: Total positive power generation by the paretic limb and distribution of power generation, measured as ankle-to-hip relative power contribution (A-H ratio), for **(A)** speed-based (Slow vs Fast) and **(B)** propulsion asymmetry-based (Moderate-Severe vs Mild) classification groups. Healthy control data provided for comparison. Abbreviation: mod-sv = Moderate-Severe; norm = young healthy control

#### Propulsion-based classifier

As expected, when compared to healthy controls, both the mild and moderate-severe propulsion asymmetry groups had significantly lower total positive power generation by their paretic limbs (p<0.001). However, and converse to the findings of the speed-based classification, whereas total positive power generation was not significantly different between the mild and moderate-severe asymmetry groups (p=0.31), paretic joint power distribution varied across the groups (p<0.001) (see **Table 2**). Whereas the A-H ratio was not significantly different across the healthy control (0.56 ± 0.26 A-H ratio) and mild asymmetry (0.57 ± 0.47 A-H ratio) groups (p=0.81), a 3x larger A-H ratio was observed in the mild asymmetry (0.57 ± 0.47 A-H ratio) versus moderate-severe asymmetry (0.19 ± 0.19 A-H ratio) groups. A similar ∼3x larger A-H ratio was observed in the healthy control group (0.56 ± 0.26 A-H ratio) compared to the moderate-severe asymmetry group (**Table 2**, **Figure 3B**).

**Table 2:** Positive power generation across joints based on propulsion classification.

| | Mod-Severe<br>Asymmetry<br>($< 31.5\%$) | Mild<br>Asymmetry<br>($> 31.5\%$) | Healthy<br>control | K-W<br>p-value | Post-hoc<br>sig pair |
| --- | --- | --- | --- | --- | --- |
| N | 14 | 17 | 12 | - | - |
| Pp (%) | 22.10 | 41.71 | 48.81 | $< 1E-3^*$ | 1-2,2-3, |
|  |  |  |  |  | 1-3 |
| ***Paretic Limb Metrics*** |  |  |  |  |  |
| Total power (J/kg) | 0.467 (0.88) | 0.821 (0.71) | 2.416 (0.68) | <1E-3* | 2-3,1-3 |
| ankle power (%) | 13.2 (11.5) | 25.2 (20.1) | 26.9 (7.1) | <1E-3* | 1-2,1-3 |
| knee power (%) | 22.1 (10.9) | 22.1 (13.8) | 25.1 (5.5) | 0.75 | - |
| hip power (%) | 61.3 (17.8) | 49.2 (22.4) | 48.5 (9.7) | 0.03* | 1-2, 1-3 |
| A/H ratio | 0.19 (0.19) | 0.57 (0.47) | 0.56 (0.26) | <1E-3* | 1-2,1-3 |
| ***Non-Paretic Limb Metrics*** |  |  |  |  |  |
| Total power (J/kg) | 0.779 (0.95) | 0.820 (0.82) | 2.416 (0.68) | <1E-3* | 2-3,1-3 |
| ankle power (%) | 28.0 (19.4) | 28.7 (8.1) | 26.9 (7.1) | 0.58 | - |
| knee power (%) | 13.4 (6.8) | 16.6 (14.6) | 25.1 (5.5) | <1E-3* | 2-3,1-3 |
| hip power (%) | 51.7 (17.1) | 54.6 (16.2) | 48.5 (9.7) | 0.25 | - |
| A/H ratio | 0.52 (0.51) | 0.61 (0.43) | 0.56 (0.26) | 0.70 | - |

Mod-Severe asymmetry group = moderate-severe asymmetry group (< 31.5%); mild asymmetry group = >31.5%; Median (interquartile range) reported; K-W test = Kruskal Wallis test; post-hoc significant pairs: 1-2 = moderate-severe vs. mild asymmetrical, 2-3 = mild asymmetrical vs. healthy control, 1-3 = moderate-severe asymmetrical vs. healthy control. \* $p < 0.05$

In the non-paretic limb analyses, both the mild and moderate-severe asymmetry groups had significantly lower total positive power generation by the non-paretic limb compared to healthy controls (p<0.001). The mild and moderate-severe groups were not significantly different (p=0.49). Moreover, the A-H ratio was not significantly different across groups (p=0.70) (see **Table 2**)

#### Speed and Propulsion: Independent predictors of paretic power generation impairments

To examine the independent associations of walking speed and propulsion asymmetry to different power generation impairments (i.e., total positive power and paretic limb power distribution) we evaluated two multiple linear regression models testing the walking speed and propulsion asymmetry classifiers as predictors. Variance in the total positive power outcome (R^2^ = 0.46, F (2,28) =11.7, p<0.001) was explained by only the speed classifier (β=+0.67, p<0.001), not the propulsion classifier (β=+0.02, p=0.92). The interaction between the speed and propulsion classifiers was not significant (p>0.05). In contrast, variance in the ankle-to-hip power distribution ratio (R^2^ = 0.37, F (2,28) =8.1, p=0.002) was explained by only the propulsion classifier (β=+0.61, p<0.001), not the speed classifier (β=-0.17, p=0.27). The interaction between the speed and propulsion classifiers was not significant (p>0.05).

### Metabolic cost of walking

Of the 24 individuals who completed metabolic cost of walking assessments, 9 were classified as slow and the remaining 15 were classified as fast based on the speed-based classification cut-off of 0.80 m/s. Normative metabolic averages from for an age-and speed-matched cohort were used for this analysis [Berryman 2012]. Post-stroke walkers in both slow and fast groups were metabolically different from each other and the healthy normative control [Berryman 2012] (p<0.001), with higher metabolic cost of walking compared to healthy normative controls. Specifically, within the post-stroke cohort, slower individuals had an 83.2% higher metabolic cost of walking (Δ=0.198 mL/kg/m, p<0.001), compared to faster individuals and 211.4% higher metabolic cost of walking (Δ=0.296 mL/kg/m, p<0.001) compared to the healthy normative control. Moreover, faster individuals also had a 70.0% higher metabolic cost of walking (Δ=0.098 mL/kg/m, p<0.001) compared to healthy normative control (**Figure 4**).

**Figure 4:**
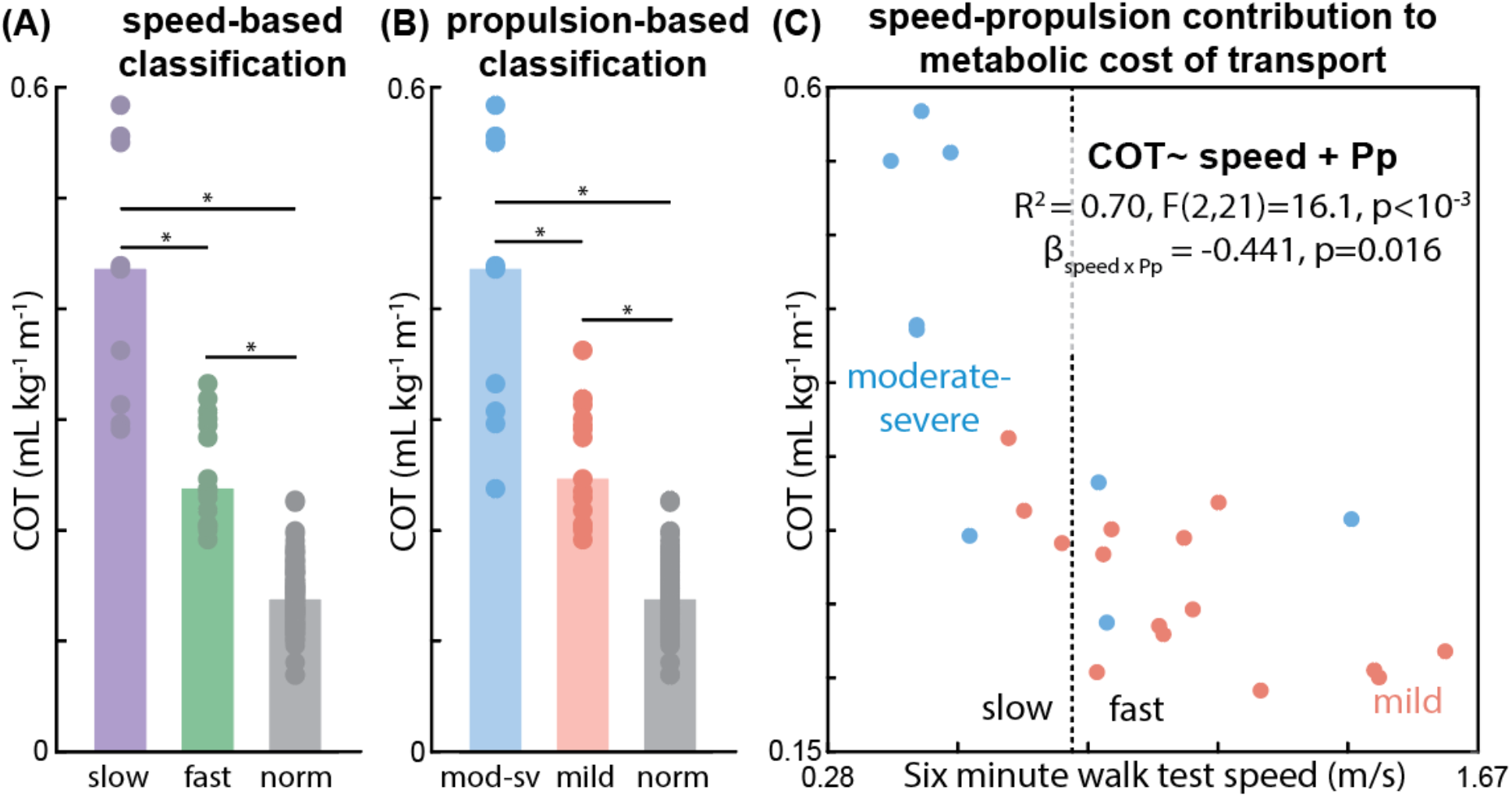
Metabolic cost of transport across speed and propulsion-based groups. left panel: speed-based classifier (Slow vs. Fast), middle panel: propulsion-based classifier (moderate-severe vs. Mild), right panel: multiple regression estimating cost of transport (COT) by speed and propulsion asymmetry groups. Abbreviation: mod-sv = Moderate-Severe; norm = healthy normative control

Based on the propulsion-based classifier, 9 were classified as having moderate-severe propulsion asymmetry and the remaining 15 were classified as having mild propulsion asymmetry. Post-stroke walkers in both moderate-severe and mild asymmetry groups were metabolically different from each other and healthy normative controls [Berryman 2012] (p<0.001), with both groups showing a higher cost of walking compared to healthy normative controls. Specifically, the moderate-severe asymmetric group had a 77.2% higher metabolic cost of walking (Δ=0.190 mL/kg/m, p=0.002), compared to the mild asymmetric group and a 211.4% higher metabolic cost of walking (Δ=0.296 mL/kg/m, p<0.001) compared to the healthy normative controls. Moreover, the mild asymmetric group also had a 75.7% higher metabolic cost of walking (Δ=0.106 mL/kg/m, p<0.001) compared to the healthy normative controls **(Figure 4)**.

Given that both the speed and propulsion classifiers had a significant influence on the metabolic cost of walking, we evaluated multiple linear regression with speed and propulsion classifiers as predictors of the metabolic cost of walking. The metabolic cost of walking was significantly explained by the interaction (β=-0.441, p=0.016) of the speed and propulsion classifiers (R^2^ = 0.70, F (2,21) =16.1, p<0.001) **(Figure 4)**.

### Ambulatory Function Measures

Fifty-seven individuals post-stroke completed assessments of the 6MWT, CWS, and propulsion asymmetry. FGA scores were available for 45 individuals, while TUG scores were available for 37 individuals. Based on the speed-based classifier, 28 out of 57 subjects, 21 out of 45 subjects, and 19 out of 37 subjects were classified as slow walkers in the CWS, FGA, and TUG datasets, respectively. Similarly, based on the propulsion-based classifier, 27 out of 57 subjects, 22 out of 45 subjects, and 16 out of 37 subjects were classified as having moderate-severe asymmetry in the CWS, FGA, and TUG datasets, respectively.

#### Speed-based classifier

Individuals post-stroke in both the slow and fast groups were functionally different from each other and healthy normative control [Morbach 2025, Bohannon 2011, Walker 2007, Bohannon 2006] (p<0.001), with both groups showing lower comfortable walking speed, lower FGA, and lower TUG scores compared to healthy normative controls. Moreover, the slow group walked 48% slower (Δ=0.47 m/s, p<0.001), had 50% lower FGA scores (Δ=10pts, p<0.001), and 81.7% higher TUG times (Δ=8.5 sec, p<0.001) compared to the fast group, and were 62.7% slower (Δ=0.84 m/s, p<0.001), had 63.1% lower FGA scores (Δ=17.1pts, p<0.001), and had 133.3% higher TUG times (Δ=10.8 sec, p<0.001) compared to healthy normative controls. Moreover, the fast group also walked 32.8% slower (Δ=0.44 m/s, p<0.001), had 26.2% lower FGA scores (Δ=7.1pts, p<0.001), and had 28.4% higher TUG times (Δ=2.3 sec, p<0.001) compared to healthy normative controls. Individuals post-stroke in both the slow and fast groups had lower propulsion asymmetry compared to healthy normative controls (p<0.001); however no significant differences in propulsion asymmetry were observed across groups (p>0.05) **(Figure 5A)**.

**Figure 5:**
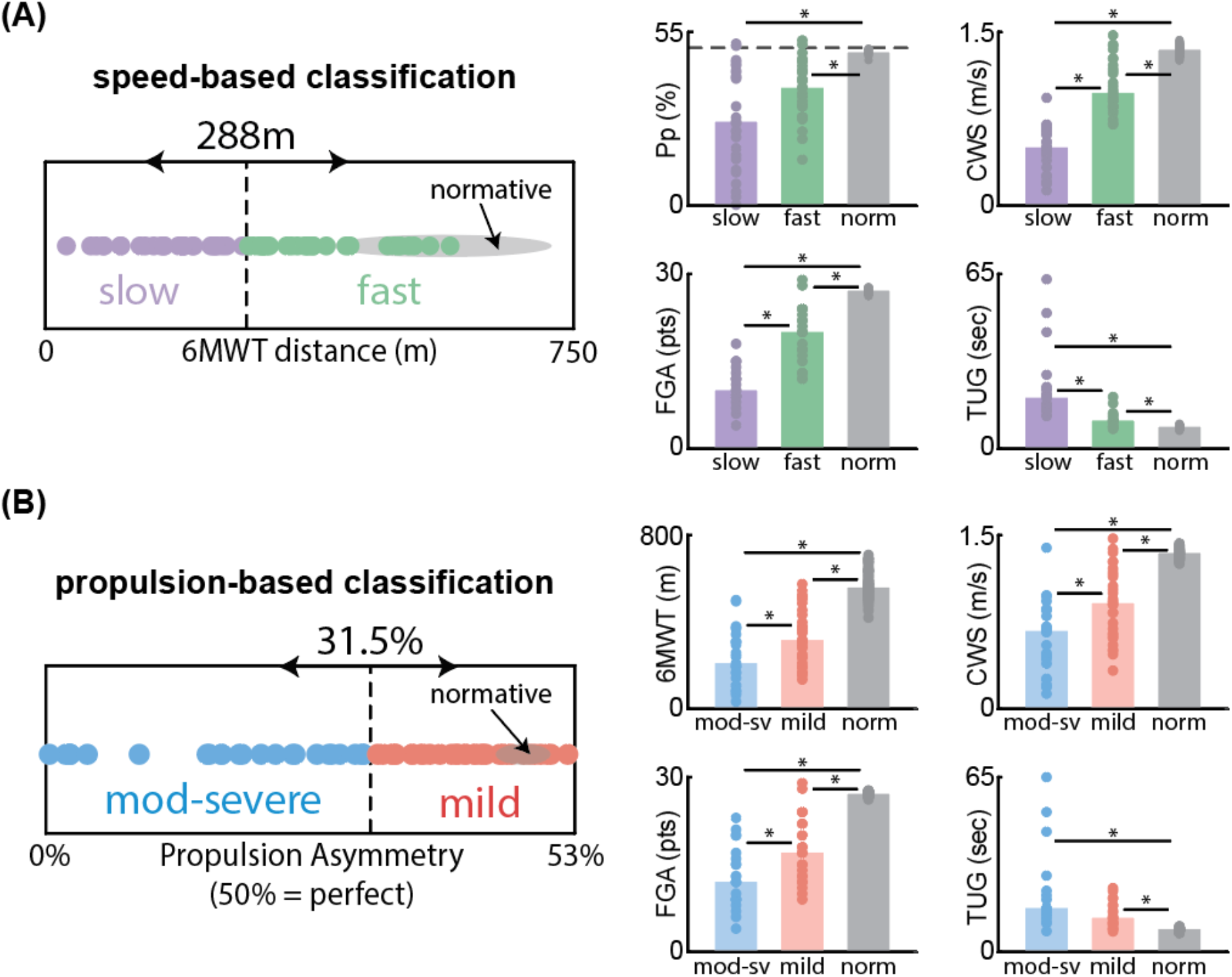
Ambulatory function abilities across. **(A)** speed-based classifier (Slow vs. Fast), **(B)** propulsion-based classifier (Moderate-severe vs. Mild), and healthy normative data. Abbreviation: mod-sv = Moderate-Severe; norm = healthy normative control

#### Propulsion-based classifier

Individuals poststroke in both the mild and moderate-severe propulsion asymmetry groups were functionally different from each other and healthy normative controls [Morbach 2025, Bohannon 2011, Walker 2007, Bohannon 2006] (p<0.001), with both groups showing lower 6MWT distances, lower comfortable walking speeds, lower FGA scores, and higher TUG times compared to healthy normative controls. Specifically, the moderate-severe asymmetry group walked 34% less distance (Δ=107m, p=0.006), 26% slower (Δ=0.24 m/s, p=0.002), and had 29% lower FGA scores (Δ=5pts, p=0.006) compared to the mild asymmetry group, and walked 62% less distance (Δ=348 m, p<0.001), 50% slower (Δ=0.67 m/s, p<0.001), and had 55% lower FGA scores (Δ=15pts, p<0.001) and 94% higher TUG times (Δ=7.6 sec, p<0.001) compared to healthy normative controls. Moreover, the mild asymmetric group also walked 43% less distance (Δ=241 m, p<0.001), 32% slower (Δ=0.43 m/s, p<0.001), and had 37% lower FGA scores (Δ=10pts, p<0.001) and 51% higher TUG times (Δ=4.1 sec, p<0.001) compared to healthy normative controls **(Figure 5B)**.

#### Speed and Propulsion contribution to general mobility

Given that both speed and propulsion classifiers had a significant influence on the FGA and TUG, we conducted a multiple linear regression with speed and propulsion asymmetry classifiers as predictors. FGA scores were significantly explained by both speed and propulsion classifiers (R2 = 0.55, F (2,44) =26, p<0.001). The relative contribution of the speed classifier was nearly three times larger (β=+0.65, p<0.001) to the contribution of the propulsion classifier (β=+0.22, p=0.048). The interaction between the speed and propulsion classifiers was not significant (p>0.05). TUG times were significantly explained by only the speed classifier, and not the propulsion classifier (R2 = 0.34, F (2,36) =8.6, p<0.001). The relative contribution of the speed classifier (β=-0.45, p=0.006) was nearly two times larger than the contribution of the propulsion classifier (β=-0.23, p=0.132). The interaction between the two classifiers was not significant (p>0.05).

## Discussion

In this paper, we demonstrate that classifying individuals with poststroke hemiparesis on the basis of their walking speed and propulsion asymmetry produces subgroups with distinct functional characteristics and differences in underlying biomechanical power generation mechanisms. Both slower individuals and individuals with moderate-to-severe propulsion asymmetry exhibited reduced functional performance and a higher metabolic cost of walking. However, whereas the speed-based classification revealed that slower individuals had lower total power generation driven by a proportionate reduction in power across the paretic joints, the propulsion-based classification revealed that individuals with moderate-severe asymmetry have a marked distal-to-proximal redistribution of joint power without impairment in total power generation.

Movement is considered a window into health and disease. Post-stroke movement impairments can vary widely, with hemiparesis leading to a fundamentally different gait pattern than is observed in healthy walking. Indeed, the unilateral nature of hemiparesis contributes to highly inefficient gait asymmetries that go beyond slow walking, highlighting the need for data-driven rehabilitation strategies that tailor interventions to specific gait deficits. In this paper, we demonstrate the potential for treatment-based classification of individuals post-stroke based on clinically-derived cutoffs of walking speed and propulsion asymmetry. That is, the speed- and propulsion-based classifiers presented in this paper identify distinct movement phenotypes that can guide intervention selection.

The emergence of these movement phenotypes intuitively follows the combination of two ideas: (1) that our movements tend towards locally “energy-efficient” strategies [Ralston 1958, Finley 2013] and (2) patients with similar deficits will naturally converge on similar energy-efficient strategies. For instance, individuals who struggle to access or control power generation from the distal muscles of the paretic limb may compensate by increasing reliance on more proximal muscles. Indeed, a common compensatory strategy known as “hip hiking” results from dropfoot. In this example, hip hiking is a secondary deviation to a primary dropfoot impairment. And, in the context of this paper, increasing hip power can compensate for propulsion deficits [Chen 2005, Kuo 2010, Farris 2015]. Recognizing the primary deviations that these strategies compensate for (i.e., the underlying biomechanical mechanisms) may allow for tailored rehabilitation strategies aimed at specific modifiable deficits rather than treating the observable symptom. In this paper, we show that speed- and propulsion-based phenotypes are distinct and are associated with unique profiles of biomechanical, metabolic, and functional characteristics.

### Speed-based phenotypes as a diagnostic biomarker

Walking speed has been described as the “sixth vital sign” [Fritz 2009] due to its correlation with multiple health metrics, including functional ability [Fulk 2010, Grau-Pellicer 2019, Fulk 2017], balance confidence [Middleton 2017], and walking efficiency [Awad 2019]. It is also a predictive biomarker of future health status [Manini 2006], functional declines [Grau-Pellicer 2019, Middleton 2015], and potential rehabilitation pathways [Dickstein 2008]. Its ease of measurement further enhances its clinical utility as a diagnostic biomarker. Here, we reaffirm the value of speed-based phenotypes in hemiparetic walking assessment. We find that slower walkers tend to have lower functional outcomes and higher metabolic costs compared to faster walkers, aligning with the contemporary understanding that faster walking speeds generally correlate with improved outcomes [Dickstein 2008]. However, caution is warranted; gait modifications that increase speed through compensatory mechanisms may lead to adverse effects over time [Bowden 2012b].

Biomechanically, we observe that slower individuals generate less total power, with this reduction proportionately distributed across the ankle, knee, and hip on both the paretic and non-paretic limbs. Interestingly, the ratio of power generation within each limb (i.e., the relative contributions of the ankle and hip muscles) is similar to that of healthy individuals, suggesting that slow walking speed alone does not define an altered power generation strategy after stroke. Consequently, rehabilitation approaches aimed at adjusting relative joint contributions may not find speed-based classifications useful. Both groups within the speed-based phenotype could benefit from general strength, endurance, and cardiovascular training, particularly through interventions targeting both limbs, such as treadmill walking.

### Propulsion-based phenotypes as a necessary and complementary diagnostic biomarker

Propulsion asymmetry has recently gained attention as a potential biomarker of hemiparetic severity in individuals post-stroke [Bowden 2006, Roelker 2019, Awad 2020]. When used alongside walking speed, it may help delineate unique gait phenotypes. Similar to the speed-based groups, both propulsion asymmetry groups demonstrated reduced ambulatory function and higher metabolic costs of walking relative to healthy controls. However, our findings reveal a notable distinction in joint power distribution between mild and moderate-severe asymmetrical walkers. While both groups demonstrate a similar capacity for total power generation by the paretic limb, individuals with moderate-severe asymmetry exhibit a pronounced shift from ankle to hip joint power generation. This distal-to-proximal redistribution of power is unique to the moderate-severe propulsion phenotype and has been observed in individuals experiencing motor fatigue [Kao 2023], possibly contributing to the increased metabolic cost associated with asymmetrical walking. Distal muscles, such as ankle plantarflexors, exhibit greater efficiency in generating positive power compared to proximal hip muscles, due to the presence of long tendons that facilitate energy redirection [Sawicki 2009, Kao 2023]. Thus, to achieve sustainable improvements in walking speed through non-compensatory pathways, rehabilitation for individuals with moderate-severe propulsion asymmetry should prioritize restoring distal power generation assuming there is latent capacity to improve. Together, our findings support the hypothesis that rehabilitation efforts focusing on biomechanical improvements in propulsion asymmetry can elevate outcomes in terms of walking speed, metabolic efficiency, and overall function. For instance, we have shown that propulsion-targeted gait training with soft robotic exosuits results in up to a 14% improvement in long-distance walking performance concurrent with a meaningful reduction in propulsion asymmetry [Porciuncula 2021].

### Implication for gait rehabilitation approaches

While increasing walking speed has traditionally been a primary focus in post-stroke rehabilitation, propulsion-based phenotypes offer a nuanced perspective that may enhance clinical decision-making. The observed distal-to-proximal redistribution of joint power in patients with moderate-severe propulsion asymmetry suggests that simply increasing speed may not yield long-term improvements. Without targeting joint-specific interventions, any speed gains may result from compensatory strategies, hindering sustainable progress [Bowden 2012b]. Our findings indicate that interventions like soft robotic ankle exosuits or plantarflexor functional electrical stimulation (FES) could be particularly beneficial for individuals with moderate-severe propulsion asymmetry, as these approaches target the specific biomechanical deficits by supplementing ankle power. Alternatively, rehabilitative approaches that resist hip-based compensation may also be helpful as reengaging the ankle might be the path of least resistance. In contrast, non-specific interventions, such as high-intensity treadmill walking, may be more effective for individuals with mild propulsion asymmetry, who, although slower than neurotypical individuals, do not exhibit intrinsic deficits in power generation strategies.

### Toward Precision Rehabilitation through Phenotype-Specific Interventions

A significant challenge in rehabilitation research is in matching interventions to the appropriate sub-populations; the result is mixed intervention outcomes with attenuated average efficacy. By identifying poststroke gait phenotypes associated with unique patterns of functional and biomechanical impairments, we can determine better which individuals might respond to specific interventions, thus advancing precision rehabilitation. Identifying speed-based phenotypes is readily implementable in clinical settings using simple tools like a stopwatch and measuring wheel or a single inertial measurement unit (IMU) that can measure walking speed [ArumukhomRevi 2021]. Unfortunately, propulsion-based phenotyping remains predominantly laboratory-bound. Our previous work has shown that propulsion can be estimated in a lab setting using three IMUs with subject-specific calibration [ArumukhomRevi 2020]. However, further research is needed to adapt this approach for routine clinical use. Regular assessment of propulsion asymmetry could help clinicians identify candidates for targeted interventions, moving beyond speed assessments alone to improve patient outcomes.

## Limitation and future work

There are several important limitations to consider in this study. Firstly, the use of normalized values (such as asymmetry or percent power) in this study, while valuable for comparing across individuals, may have a higher signal-to-noise ratio. This is particularly relevant for individuals with low propulsion in both limbs, where they may appear to have moderately symmetrical gait if the non-paretic limb’s propulsion magnitude is only slightly higher than that of the paretic limb. Future studies should consider incorporating absolute propulsion values in addition to asymmetry to address this issue.

Age-matched control – healthy aging affects the gait biomechanics. In fact, recent studies have shown that older adults use a hip-dominate power generation strategies compared to younger adults and subsequently have a higher metabolic cost of walking [Boyer 2023, Boyer 2017]. However, since all our post-stroke groups have a similar mean age, the effect of speed or propulsion asymmetry deficit further promotes this maladaptive gait pattern. Future studies should further examine the independent effects of age.

The 6MWT was conducted on a 27m oval track instead of the standard 30m walkway, which was necessary for instrumentation purposes. However, it is important to note that this modification may have influenced individuals’ performance, particularly due to the absence of turns. Future studies could compare the results between the modified track and the standard walkway to assess any potential significant differences.

Though average positive power generation across the gait cycle was used in this paper, the generation of power at different parts of the gait cycle is important. For example, hip power generated at stance is associated with propulsion, while the same power generated at swing is associated with limb advancement [Farris 2015]. Future work should further investigate the power generation difference observed in the propulsion-phenotypes.

While it is evident that propulsion asymmetry is a valuable measure for distinguishing individual biomechanical differences, the measurement of propulsion asymmetry is currently not easily accessible in a clinical setting. Previous studies have attempted to estimate propulsion using wearable sensors [ArumukhomRevi 2020], but they are not yet ready for clinical use. Future research should focus on developing clinically accessible methods to estimate propulsion asymmetry or its proxy measures to fully utilize this knowledge.

## Conclusion

This study underscores the critical and complementary roles that measurements of walking speed and propulsion asymmetry play in our understanding of post-stroke gait dysfunction. Our findings reveal that propulsion-based phenotypes are distinct from speed-based phenotypes. Though both classification approaches reveal phenotypes associated with impaired ambulatory function and an inefficient gait, unique power generation impairments that require different targeted gait interventions distinctively emerge within each phenotype. Indeed, whereas slow walking indicates reduced paretic limb power, a distal-to-proximal redistribution of joint power was observed only in those with moderate-to-severe propulsion asymmetry. These findings motivate the study of phenotype-guided gait interventions and the development of point-of-care diagnostic technologies that enable the dual measurement of walking speed and propulsion asymmetry as distinct diagnostic biomarkers of modifiable gait impairments. When applied to the management of the highly heterogeneous gait impairments observed after stroke, this approach has the potential to advance precision rehabilitation and optimize treatment outcomes.

## Data Availability

All data produced in the present study are available upon reasonable request to the authors

## Acknowledgement

We would like to thank Ms. Lillian Ribeirinha-Braga for all the help with participant coordination; Mr. Willie Swift, Mr. Victor Dos Reis, and Ms. Jessie Spada for help with data processing; Dr. Franchino Porciuncula for the moral support, and all our study participants for generously sharing their time.

## Author Contributions

DAR and LNA conceived the study design and experimental methods; DAR and JS conducted the experiment on the *Biomechanics Day*; DAR and AC conducted the experiments on the *Metabolic Day*; DAR and LNA conceived the data analysis approach; DAR conducted the data analysis; MGB, SDR and TE provide substantial feedback on data analysis and/or results interpretation. DAR, MGB and LNA prepared the manuscript; all authors revised and approved the final manuscript.

## Ethics

This study was approved by the Boston University Charles River Campus Institutional Review Board (protocol #4440), and all procedures were conducted in accordance with the Declaration of Helsinki. Written informed consent was obtained from all participants prior to enrollment.

## Appendix A: Classification of mild vs. severe hemiparetic propulsion asymmetry

### Background

In Bowden 2006 [Bowden 2006], the Brunnstrom stages of recovery were used to anchor the propulsion deficits observed in individuals post-stroke. Individuals with a Brunnstrom stage of 3 or less were identified as having a mild hemiparetic deficit, while individuals with Brunnstrom stage of 4 or 5 were identified as having a moderate hemiparetic deficit (36%). Individuals with Brunnstrom stage of 6 were identified as having a severe hemiparetic deficit. Unfortunately, this manuscript does not conduct any formal analysis to determine a cutoff in propulsion asymmetry that can be used to specific deficit levels (mild, moderate, and/or severe). Other studies that aim to identify individuals between mild and severe hemiparetic deficits tend to use the 36% threshold, which is the average of the moderate group and is importantly not appropriate for this usage. Therefore, the goal of this appendix is to provide a threshold in propulsion asymmetry that can distinguished individuals as having mild vs. severe hemiparetic deficits.

### Methods

Forty-seven individuals from the original paper [Bowden 2006] were used for this analysis. Propulsion asymmetry was defined as paretic limb propulsion divided by the sum of paretic and non-paretic limb propulsion, such that 50% represents perfect symmetry. If paretic limb propulsion was higher than the non-paretic limb, the propulsion asymmetry was defined as non-paretic limb propulsion divided by the sum of paretic and non-paretic limb propulsion. For this analysis, propulsion asymmetry is always bounded between 0 and 50%. We distinguished individuals with an Brunnstrom stage of recovery score less than or equal to four as having mild hemiparetic deficit, and individuals with an Brunnstrom stage of recovery score greater than four as having severe hemiparetic deficit. An ROC analysis was performed to identify the cutoff in propulsion asymmetry that balances accuracy, sensitivity and specificity to distingue mild vs. severe hemiparetic deficit.

### Results and Discussion

Based on the Brunnstrom stage of recovery score, we identified 24 individuals as having a severe hemiparetic deficit (Brunnstrom score > 4) and 23 individuals as having a mild hemiparetic deficit (Brunnstrom score <= 4). A propulsion asymmetry threshold of 31.5% can distinguish individuals with severe vs. mild hemiparetic deficit, with an accuracy of 83.0%, sensitivity of 82.6%, and specificity of 83.3% (Figure A1). Whereas the commonly used propulsion asymmetry threshold of 36% can distinguish individuals with severe vs. mild hemiparetic deficit, with an accuracy of 80.9%, sensitivity of 73.9%, and specificity of 87.5% (Figure A1).

**Figure A1:**
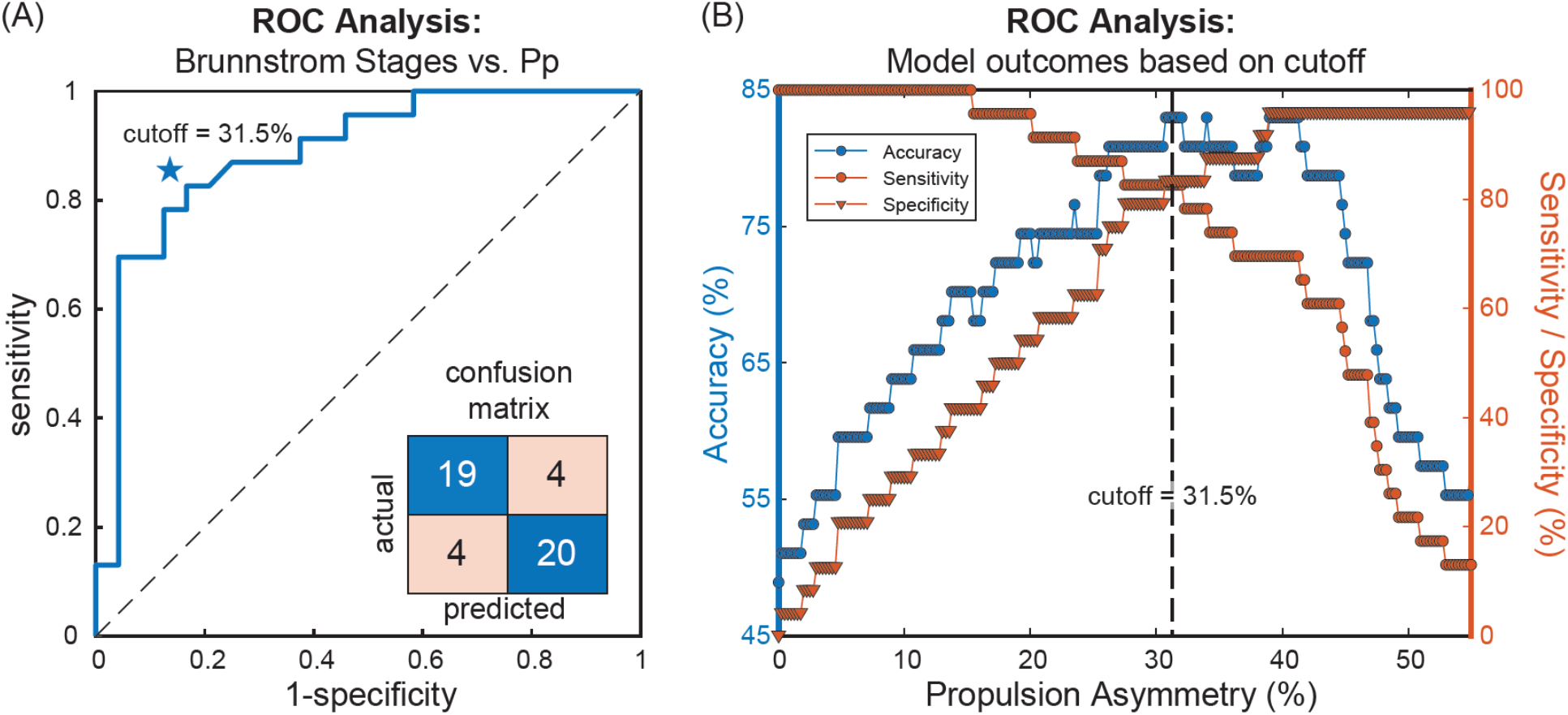
**(A)** ROC analysis of Brannstrom stages vs. propulsion asymmetry used to determine cutoff in Pp that can distinguish mild vs. severe hemiparesis and confusion matrix results **(B)** Accuracy, sensitivity and specificity of model based on cutoff.

Here we provide a threshold for propulsion asymmetry that can be used to distinguish between mild vs. severe post-stroke hemiparesis based on the Brunnstrom stage of recovery score that balances accuracy, sensitivity and specificity. Future studies can now utilize this threshold of 31.5% instead of the currently used 36% threshold to characterize post-stroke cohorts better.

## References

1. Alexander RM. Elastic Mechanisms in Animal Movement. Cambridge University Press; 1988.

2. Arumukhom Revi D, Alvarez AM, Walsh CJ, De Rossi SMM, Awad LN. Indirect measurement of anterior-posterior ground reaction forces using a minimal set of wearable inertial sensors: from healthy to hemiparetic walking. J Neuroeng Rehabil. 2020;17(1):82.

3. Arumukhom Revi D, De Rossi SMM, Walsh CJ, Awad LN. Estimation of walking speed and its spatiotemporal determinants using a single inertial sensor worn on the thigh: from healthy to hemiparetic walking. Sensors (Basel). 2021;21(21):6976.

4. Awad LN, Reisman DS, Pohlig RT, Binder-Macleod SA. Reducing the cost of transport and increasing walking distance after stroke. Neurorehabil Neural Repair. 2016;30(7):661–670.

5. Awad L, Reisman D, Binder-Macleod S. Distance-induced changes in walking speed after stroke: relationship to community walking activity. J Neurol Phys Ther. 2019;43(4):220–223.

6. Awad LN, Lewek MD, Kesar TM, Franz JR, Bowden MG. These legs were made for propulsion: advancing the diagnosis and treatment of post-stroke propulsion deficits. J Neuroeng Rehabil. 2020;17(1):139.

7. Barroso FO, Torricelli D, Molina-Rueda F, et al. Combining muscle synergies and biomechanical analysis to assess gait in stroke patients. J Biomech. 2017;63:98–103.

8. Berryman N, Gayda M, Nigam A, Juneau M, Bherer L, Bosquet L. Comparison of the metabolic energy cost of overground and treadmill walking in older adults. Eur J Appl Physiol. 2012;112(5):1613–1620.

9. Bohannon RW. Reference values for the timed up and go test: a descriptive meta-analysis. J Geriatr Phys Ther. 2006;29(2):64–68.

10. Bohannon RW, Andrews AW. Normal walking speed: a descriptive meta-analysis. Physiotherapy. 2011;97(3):182–189.

11. Bowden MG, Balasubramanian CK, Neptune RR, Kautz SA. Anterior-posterior ground reaction forces as a measure of paretic leg contribution in hemiparetic walking. Stroke. 2006;37(3):872–876.

12. Bowden MG, Balasubramanian CK, Behrman AL, Kautz SA. Validation of a speed-based classification system using quantitative measures of walking performance poststroke. Neurorehabil Neural Repair. 2008;22(6):672–675.

13. Bowden MG, Behrman AL, Woodbury M, Gregory CM, Velozo CA, Kautz SA. Advancing measurement of locomotor rehabilitation outcomes to optimize interventions and differentiate between recovery versus compensation. J Neurol Phys Ther. 2012;36(1):38–44.

14. Bowden MG, Embry AE, Perry LA, Duncan PW. Rehabilitation of walking after stroke. Curr Treat Options Neurol. 2012;14(6):521–530.

15. Boyer KA, Johnson RT, Banks JJ, Jewell C, Hafer JF. Systematic review and meta-analysis of gait mechanics in young and older adults. Exp Gerontol. 2017;95:63–70.

16. Boyer KA, Hayes KL, Umberger BR, et al. Age-related changes in gait biomechanics and their impact on the metabolic cost of walking: report from a National Institute on Aging workshop. Exp Gerontol. 2023;173:112102.

17. Chen G, Patten C, Kothari DH, Zajac FE. Gait differences between individuals with post-stroke hemiparesis and non-disabled controls at matched speeds. Gait Posture. 2005;22(1):51–56.

18. Cofré LE, Lythgo N, Morgan D, Galea MP. Aging modifies joint power and work when gait speeds are matched. Gait Posture. 2011;33(3):484–489.

19. Combs SA, Van Puymbroeck M, Altenburger PA, Miller KK, Dierks TA, Schmid AA. Is walking faster or walking farther more important to persons with chronic stroke? Disabil Rehabil. 2013;35(10):860–867.

20. Dickstein R. Rehabilitation of gait speed after stroke: a critical review of intervention approaches. Neurorehabil Neural Repair. 2008;22(6):649–660.

21. Donelan JM, Kram R, Kuo AD. Mechanical and metabolic determinants of the preferred step width in human walking. Proc R Soc Lond B Biol Sci. 2001;268(1480):1985–1992.

22. Donelan JM, Kram R, Kuo AD. Simultaneous positive and negative external mechanical work in human walking. J Biomech. 2002;35(1):117–124.

23. Ellis RG, Howard KC, Kram R. The metabolic and mechanical costs of step time asymmetry in walking. Proc Biol Sci. 2013;280(1756):20122784.

24. Farris DJ, Sawicki GS. The mechanics and energetics of human walking and running: a joint level perspective. J R Soc Interface. 2012;9(66):110–118.

25. Farris DJ, Hampton A, Lewek MD, Sawicki GS. Revisiting the mechanics and energetics of walking in individuals with chronic hemiparesis following stroke: from individual limbs to lower limb joints. J Neuroeng Rehabil. 2015;12(1):24.

26. Feigin VL, Stark BA, Johnson CO, et al. Global, regional, and national burden of stroke and its risk factors, 1990-2019: a systematic analysis for the Global Burden of Disease Study 2019. Lancet Neurol. 2021;20(10):795–820.

27. Finley JM, Bastian AJ, Gottschall JS. Learning to be economical: the energy cost of walking tracks motor adaptation. J Physiol. 2013;591(4):1081–1095.

28. French MA, Roemmich RT, Daley K, et al. Precision rehabilitation: optimizing function, adding value to health care. Arch Phys Med Rehabil. 2022;103(6):1233–1239.

29. Fritz S, Lusardi M. White paper: “walking speed: the sixth vital sign”. J Geriatr Phys Ther. 2009;32(2):2–5.

30. Fulk GD, Reynolds C, Mondal S, Deutsch JE. Predicting home and community walking activity in people with stroke. Arch Phys Med Rehabil. 2010;91(10):1582–1586.

31. Fulk GD, He Y, Boyne P, Dunning K. Predicting home and community walking activity poststroke. Stroke. 2017;48(2):406–411.

32. Grau-Pellicer M, Chamarro-Lusar A, Medina-Casanovas J, Serdà Ferrer BC. Walking speed as a predictor of community mobility and quality of life after stroke. Top Stroke Rehabil. 2019;26(5):349–358.

33. Hsiao H, Knarr BA, Pohlig RT, Higginson JS, Binder-Macleod SA. Mechanisms used to increase peak propulsive force following 12-weeks of gait training in individuals poststroke. J Biomech. 2016;49(3):388–395.

34. Jonkers I, Delp S, Patten C. Capacity to increase walking speed is limited by impaired hip and ankle power generation in lower functioning persons post-stroke. Gait Posture. 2009;29(1):129–137.

35. Kao PC, Lomasney C, Gu Y, Clark JP, Yanco HA. Effects of induced motor fatigue on walking mechanics and energetics. J Biomech. 2023;156:111688.

36. Kuo AD, Donelan JM, Ruina A. Energetic consequences of walking like an inverted pendulum: step-to-step transitions. Exerc Sport Sci Rev. 2005;33(2):88–97.

37. Kuo AD, Donelan JM. Dynamic principles of gait and their clinical implications. Phys Ther. 2010;90(2):157–174.

38. Mahon CE, Farris DJ, Sawicki GS, Lewek MD. Individual limb mechanical analysis of gait following stroke. J Biomech. 2015;48(6):984–989.

39. Manini TM, Everhart JE, Patel KV, et al. Daily activity energy expenditure and mortality among older adults. JAMA. 2006;296(2):171–179.

40. Middleton A, Fritz SL, Lusardi M. Walking speed: the functional vital sign. J Aging Phys Act. 2015;23(2):314–322.

41. Middleton A, Braun CH, Lewek MD, Fritz SL. Balance impairment limits ability to increase walking speed in individuals with chronic stroke. Disabil Rehabil. 2017;39(5):497–502.

42. Morbach C, Moser N, Cejka V, et al. Determinants and reference values of the 6-min walk distance in the general population—results of the population-based STAAB cohort study. Clin Res Cardiol. 2025;114: 1098–1108

43. Nadeau S, Gravel D, Arsenault AB, Bourbonnais D. Plantarflexor weakness as a limiting factor of gait speed in stroke subjects and the compensating role of hip flexors. Clin Biomech (Bristol, Avon). 1999;14(2):125–135.

44. Parikh V, Slusarenko A, Spencer J, et al. Biomechanical and neural correlates of FastFES versus Fast gait training in individuals post stroke: a randomized control trial study protocol. Front Neurol. 2026;17:1792419.

45. Podsiadlo D, Richardson S. The timed “Up & Go”: a test of basic functional mobility for frail elderly persons. J Am Geriatr Soc. 1991;39(2):142–148.

46. Porciuncula F, Baker TC, Arumukhom Revi D, et al. Targeting paretic propulsion and walking speed with a soft robotic exosuit: a consideration-of-concept trial. Front Neurorobot. 2021;15:689577.

47. Porciuncula F, Arumukhom Revi D, Baker TC, et al. Effects of high-intensity gait training with and without soft robotic exosuits in people post-stroke: a development-of-concept pilot crossover trial. J Neuroeng Rehabil. 2023;20(1):148.

48. Ralston HJ. Energy-speed relation and optimal speed during level walking. Int Z Angew Physiol Einschl Arbeitsphysiol. 1958;17(4):277–283.

49. Reisman DS, Rudolph KS, Farquhar WB. Influence of speed on walking economy poststroke. Neurorehabil Neural Repair. 2009;23(6):529–534.

50. Roelker SA, Bowden MG, Kautz SA, Neptune RR. Paretic propulsion as a measure of walking performance and functional motor recovery post-stroke: a review. Gait Posture. 2019;68:6–14.

51. Sadeghi H, Allard P, Prince F, Labelle H. Symmetry and limb dominance in able-bodied gait: a review. Gait Posture. 2000;12(1):34–45.

52. Sawicki GS, Lewis CL, Ferris DP. It pays to have a spring in your step. Exerc Sport Sci Rev. 2009;37(3):130–138.

53. Swaminathan K, Porciuncula F, Park S, et al. Ankle-targeted exosuit resistance increases paretic propulsion in people post-stroke. J Neuroeng Rehabil. 2023;20(1):85.

54. Walker ML, Austin AG, Banke GM, et al. Reference group data for the Functional Gait Assessment. Phys Ther. 2007;87(11):1468–1477.

55. Wrisley DM, Marchetti GF, Kuharsky DK, Whitney SL. Reliability, internal consistency, and validity of data obtained with the Functional Gait Assessment. Phys Ther. 2004;84(10):906–918.

